# Distance matters: barriers to antenatal care and safe childbirth in a migrant population on the Thailand-Myanmar border from 2007-2015, a pregnancy cohort study

**DOI:** 10.1101/2020.11.13.20231464

**Authors:** Eric Steinbrook, Myo Chit Min, Ladda Kajeechiwa, Jacher Wiladphaingern, Moo Kho Paw, MuPawJay Pimanpanarak, Woranit Hiranloetthanyakit, Aung Myat Min, Nay Win Tun, Mary Ellen Gilder, François Nosten, Rose McGready, Daniel M. Parker

## Abstract

**Background:** Antenatal care and delivery with skilled attendants substantially improve maternal health outcomes across the pregnancy spectrum, from conception to delivery. The Sustainable Development Goals recognize the need to expand these services to all pregnant women but there is limited data on access for migrants and in post-conflict regions.

**Methods:** Using geographic information systems established for malaria elimination efforts in Kayin state, Myanmar and Tak Province, Thailand, retrospective estimates of travel distances from home villages to maternal health facilities between 2007-2015 were made. Multivariable regressions were used to assess the relationships between distance to healthcare and 1) presentation for early pregnancy care, 2) complications during pregnancy like malaria infections, and 3) eventual outcome of the pregnancy.

**Findings:** Women who delayed antenatal care until the third trimester travelled 46% farther (DR: 1.46; CI: 1.39 – 1.53) compared to women who attended in the first trimester, and those with pregnancies complicated by *Plasmodium falciparum* malaria travelled 62% farther (DR: 1.62; CI: 1.44 – 1.82) than those without *P. falciparum*. Women did not deliver with skilled birth services and were lost to follow-up travelled 45% farther (distance ratio (DR): 1.45; CI: 1.40 – 1.51) than those who followed-up to deliver with skilled birth services.

**Interpretation:** This analysis supports substantial global evidence that travel distance disrupts access to care in limited resource regions. This is the first demonstration of empirical distance impacting maternal healthcare from early pregnancy to delivery of migrants living in post-conflict Thailand-Myanmar border regions, and future interventions should provide decentralized maternal healthcare to address these barriers.

**Funding:** The Bill & Melinda Gates Foundation and the Wellcome Trust

## INTRODUCTION

For over 50 years civil war in Kayin state, Myanmar has forced hundreds of thousands to flee their homes and resettle either in the relative security of the Thai/Myanmar border region or to a third country.^1^ Peace talks began in 2013, but the legacy of the civil war continues to manifest in the region. A lack of public health infrastructure limits accessibility for those who remain in Myanmar,^1^ and language barriers and a lack of official immigration status prevents the many people who seek employment in Thailand from accessing services run by the Thai healthcare system. Although health data for this conflict-displaced population has been difficult to collect, what has been gathered indicates a high burden of poor pregnancy outcomes, mostly driven by conditions that can be reduced or prevented through routine antenatal care and skilled birth attendants.^2^

WHO-approved interventions across the pregnancy spectrum (Figure 1), from early pregnancy care during the first trimester to delivery in the presence of skilled birth attendants in the third trimester, have been shown to improve pregnancy outcomes.^3^ A growing body of literature, mainly focusing on low-income and resource-limited settings, has shown that geographic distance to maternal health facilities influences women’s use of those facilities. A systematic review of west African countries found that travel distance was a key predictor of maternal healthcare services^4^, and a prospective cohort study in rural Rwanda found that distance from delivery services significantly predicts the use of skilled birth attendants, but does not affect the number of visits women make to antenatal care facilities.^5^ The majority of evidence comes from cross-sectional studies in Cameroon, Rwanda, Ethiopia, China, Burkina Faso, and Ghana, which indicate that distance to healthcare facilities is a major determinant of the use of maternal healthcare services.^6–10^ Notably, a spatial analysis from Kenya found that distance did not always correlate with use of antenatal care, which highlights how the effect of distance on uptake of maternal healthcare can vary across a region.^11^

**Figure 1:**
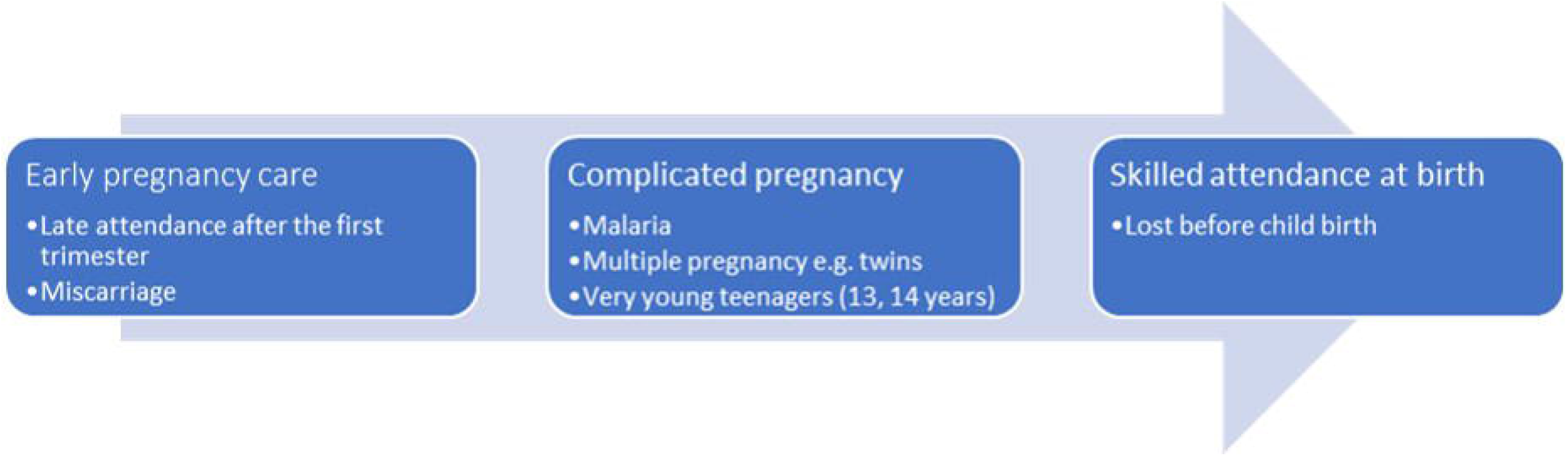
Outcomes across the pregnancy spectrum.

Distance has also been shown to correlate with delays in seeking care for pregnancy complications^12^ and receiving timely treatment for emergent conditions.^13^ Further, facility births where skilled attendants can provide emergency obstetric and newborn care are also reduced by greater distance to the health facility in low-income countries or marginalized communities^5,14^. Even a one kilometre increase in travel distance has been shown to reduce use of skilled birth attendants by 6.7%.^15^ In the Thai-Myanmar border region, however, the relationship between geographic distance, loss to follow-up, and poor pregnancy outcomes has yet to be examined.

The Shoklo Malaria Research Unit (SMRU) operates a system of antenatal clinics (ANCs) and skilled birth facilities that provide free care on the Thai-Myanmar border to marginalized populations. Many women from Myanmar, which is a low-income country, must traverse difficult terrain in order to reach these ANCs. This includes traveling on dirt roads through forested mountainous areas that can be impassable in the rainy season, rivers, and border checkpoints. In the last two decades, deforestation and the establishment of year-round roads around health facilities (Figure 2) has increased the ease of travel and likely contributed to a decrease in malaria prevalence in the areas surrounding clinics.^1^ Women attending SMRU ANCs from Thailand, which is an upper middle-income country, are predominantly undocumented, and though the roads are well maintained and SMRU provides subsidized transportation for women living far from ANC facilities there is some danger of being arrested or fined at a check-point on their way to receive care.^16^

**Figure 2:**
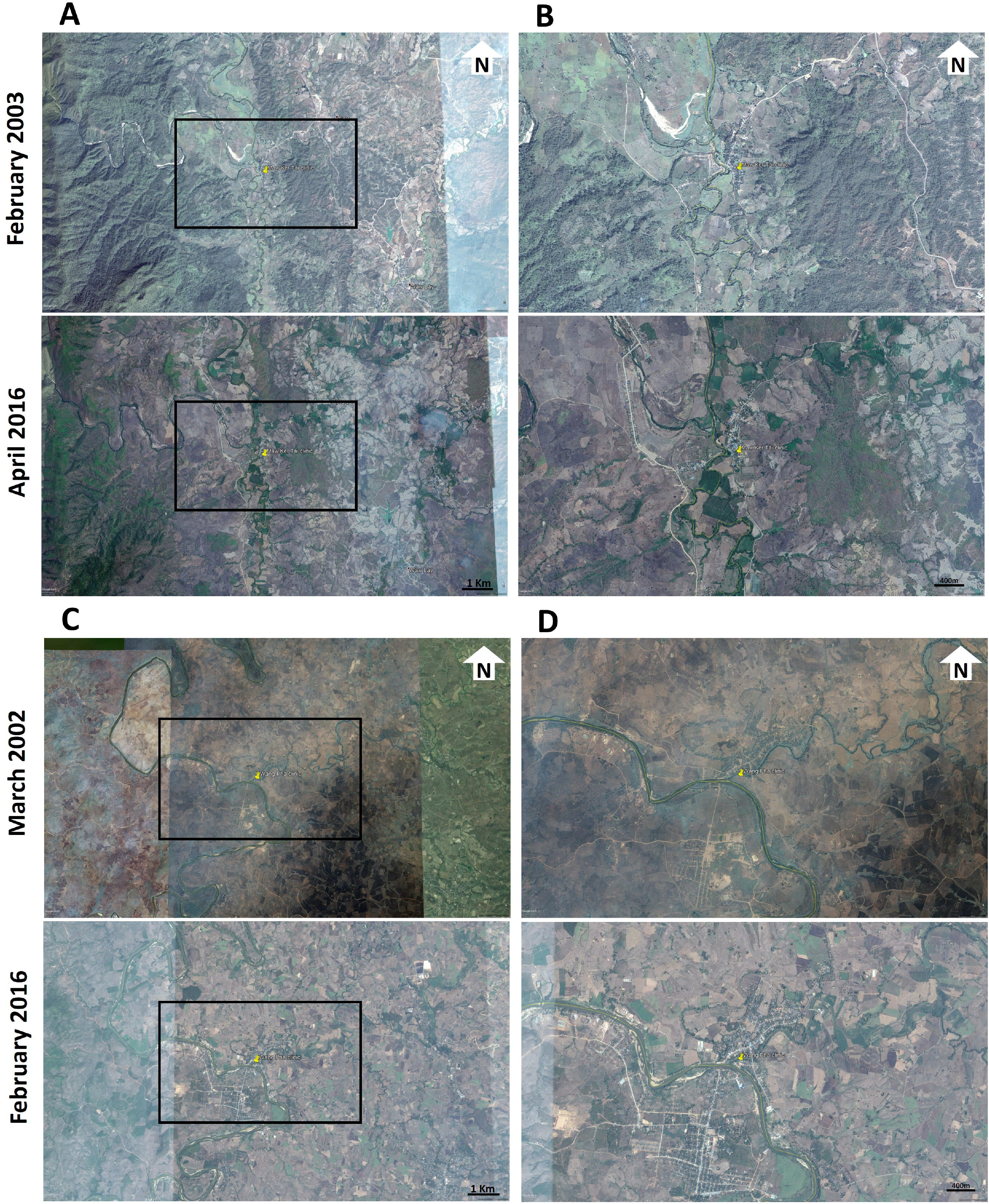
Satellite images showing environmental changes over time for two of the antenatal clinics: Mawker Tai (top panel: A and B) and Wang Pha (bottom panel: C and D) clinics. The first columns (A for Mawker Tai and C for Wang Pha) show the larger geographic area whereas the second columns (B and D) show greater detail in the immediate clinic area (zoomed areas are indicated with the black boxes in A and C). The top rows show historical images and the bottom rows show more recent images. Deforestation is apparent in apparent, especially in the Mawker Tai images and on both the Thai and Myanmar side of the international border. Infrastructure and development has also increased at both clinics, with increased year-round roads, housing, and increased agricultural fields. Images come from Google Earth.

Clinic records show that 17.4% of women who enrolled at the migrant ANCs stopped attending before giving birth and were lost to follow-up,^2^ potentially indicating that there are barriers (i.e. geographic, socio-economic, or perceptions of antenatal services) preventing women from accessing care. A previous observational study in this population observed higher loss to follow-up in women with malaria infection, first pregnancy, and younger age of the mother.^17^ A comprehensive analysis of factors influencing loss to follow-up and how those factors change from year to year has yet be completed. This manuscript investigates the relationship between travel distance and access to the full spectrum of pregnancy care. This work seeks to inform future public health interventions that address these problems, both locally and globally, where efforts to expand maternal healthcare access are ongoing.

## DATA AND METHODS

SMRU has been based on the Thai-Myanmar border for over 30 years. In response to an estimated maternal mortality in refugees of 1,000 per 100,000 live births in 1985-86, SMRU established a system of weekly ANCs to offer early detection and treatment of *Plasmodium falciparum* malaria in pregnancy.^18^ As the population of migrant workers grew in the 1990’s and 2000’s, SMRU opened four facilities on the international border formed by the Moei River, which provided a mixture of antenatal care and skilled birth services. Mawker Thai (MKT) began providing ANC and skilled birth services in 1998, Walley (WAL) and MuRu Chai (MRC) began providing ANC care in 2001, and Wang Pha (WPA) began providing ANC and skilled birth services in 2004. WAL closed operations in Jul 2010 and MRC in Dec 2012 and services were amalgamated at MKT. All services were free of charge^19^ and attendance at ANCs was voluntary.

The maternal health facilities kept antenatal medical records for each pregnancy from 1986-2015 which were de-identified and include general demographic information (patient age, gravidity, parity, home village name, and time lived at home village), antenatal care attendance information (estimated gestational age at initial presentation, miscarriages), pregnancy complication information (malaria infection with *P. vivax, P. falciparum*, or both, multiple pregnancy, very young age), and presence of skilled birth attendants at delivery (loss to follow-up and normal singleton delivery) (Figure 1).^20^

Loss to follow-up was defined as a women who enrolled at ANC but then stopped attending and did not return for delivery or postpartum care. Most women travel on foot to ANC appointments and a minority hire motorbikes or long-tractors for transportation. SMRU subsidizes transportation by car to prenatal visits for those who live on the Thai side of the border. All four facilities are built on the Thailand bank of Moei River and, depending on seasonal variations in rainwater, women use temporary bridges or boats to cross the river.

In 2014, SMRU worked to create and update a geographic information system (GIS) database for Kayin state, Myanmar and Tak province, Thailand. In Kayin State, SMRU travelled to remote villages by car, boat, or foot to obtain coordinates, which was the first systematic geographic study in the area since before World War II.^21^ In Tak province, the Tak Malaria Initiative^22^ had previously gathered village GPS coordinates, and SMRU performed an updated geographic survey in 2014. The Kayin and Tak GISs were used to link each unique pregnancy with GPS coordinates of their home village.

### Linking geospatial data to de-identified patient data

The study team used geocoding to convert place names to map coordinates. They were blinded to all information in the patient record except village names, which they matched with coordinate data in the Kayin and Tak GISs. For the portion of villages not listed in GISs (n=105/1152), ANC clinic administrators with more than 20 years of experience pinpointed villages using Google Earth software. Patient addresses weren’t consistently recorded at ANC clinics until late 2006 and the study team therefore limited this analysis to 2007 through 2015. All village names with unresolved locations were excluded from analysis, as were villages lying over 35km from the ANC given the possibility that these were mistaken village locations that did not represent actual patient experiences.

### Univariate analyses

To understand travel distance to the facilities the straight-line (Euclidian) distance between each patient’s village and the facility they attended were measured and used to calculate summary statistics and univariate analyses. The univariate analyses assessed whether travel distances differed across women based on year of childbirth, parity, age, malaria infection status, and pregnancy outcome. Travel distances across pregnancy outcomes (i.e. singleton delivery, twins, lost to follow-up, or miscarriage) was also compared. Miscarriage was defined as birth before 28 weeks gestational age based on published clinical experience of viability in this setting, where ventilatory support for newborns is not available.^17^

### Formal analysis: negative binomial regression

Negative binomial regression was used to formally analyse potential predictors (i.e. covariates) of the straight-line distance travelled to a facility. Final predictor variables (covariates) in the models included: pregnancy outcome, maternal age in years, parity, year of childbirth, and years lived at the current village (Supplementary Table 1). A sensitivity analysis was conducted to attempt to account for potential error in geocoding (described in detail in Supplementary materials).

**Table 1:** Distance from home village to health facility by year of childbirth^a^.

| **year of childbirth** | **min** | **Q1** | **median** | **Q3** | **max** | **count** |
| --- | --- | --- | --- | --- | --- | --- |
| 2007 | 0.00 | 0.89 | 1.97 | 5.47 | 33.82 | 1279 |
| 2008 | 0.00 | 0.89 | 1.97 | 6.58 | 33.82 | 1525 |
| 2009 | 0.00 | 0.89 | 2.17 | 6.96 | 30.98 | 1844 |
| 2010 | 0.00 | 0.89 | 4.72 | 8.06 | 30.62 | 1950 |
| 2011 | 0.00 | 0.89 | 6.27 | 8.70 | 33.82 | 1933 |
| 2012 | 0.00 | 0.89 | 6.41 | 9.35 | 30.98 | 1824 |
| 2013 | 0.00 | 2.17 | 6.96 | 12.87 | 31.27 | 2032 |
| 2014 | 0.00 | 3.93 | 7.52 | 15.20 | 31.27 | 2154 |
| 2015 | 0.00 | 2.17 | 7.52 | 15.44 | 33.82 | 2167 |

<sup>a</sup> WAL clinic closed in June 2010, and patients were amalgamated to MKT. MRC clinic closed in December 2012, and patients were amalgamated to MKT.

### Statistical software

Maps were created using QGIS version 3.4.9. The Python programming language (version 3.6) was used to merge geocoded home villages to the patient records. R statistical software version 3.3.2 was used for all statistical analyses.

## RESULTS

### Summary statistics and univariate analyses

There were 17,522 unique pregnancy records with complete data from SMRU’s four facilities for the study period of 2007-2015 identified. Successful linkage of 97.9% of these records (17,162/17,522) to GPS coordinates in the GIS databases was achieved. The remaining, unlinked records either did not list home villages, were recorded illegibly, or could not be located in the GIS databases and were excluded from analyses.

Figure 3 shows the estimated catchment areas for each of the four facilities across the study period (changes in catchment area over time are shown in Supplementary Figure 2). Women attending the WPA and MKT clinics travelled a longer distance to receive care compared to those attending WAL and MRC clinics. Median distance travelled increased over time as well (Table 1). At the beginning of the study period opened, most women were coming from nearby (Supplementary Table 4 and Supplementary Figure 2). Over time, women came from farther away, increases in travel distance are noted around June 2010 when WAL patients were amalgamated to MKT, and in December 2012 when MRC patients were amalgamated to MKT. In 2007-2008 women travelled a median distance of less than 2km, but for 2011-2015 the median distance increased to over 7km (Table 1).

**Figure 3:**
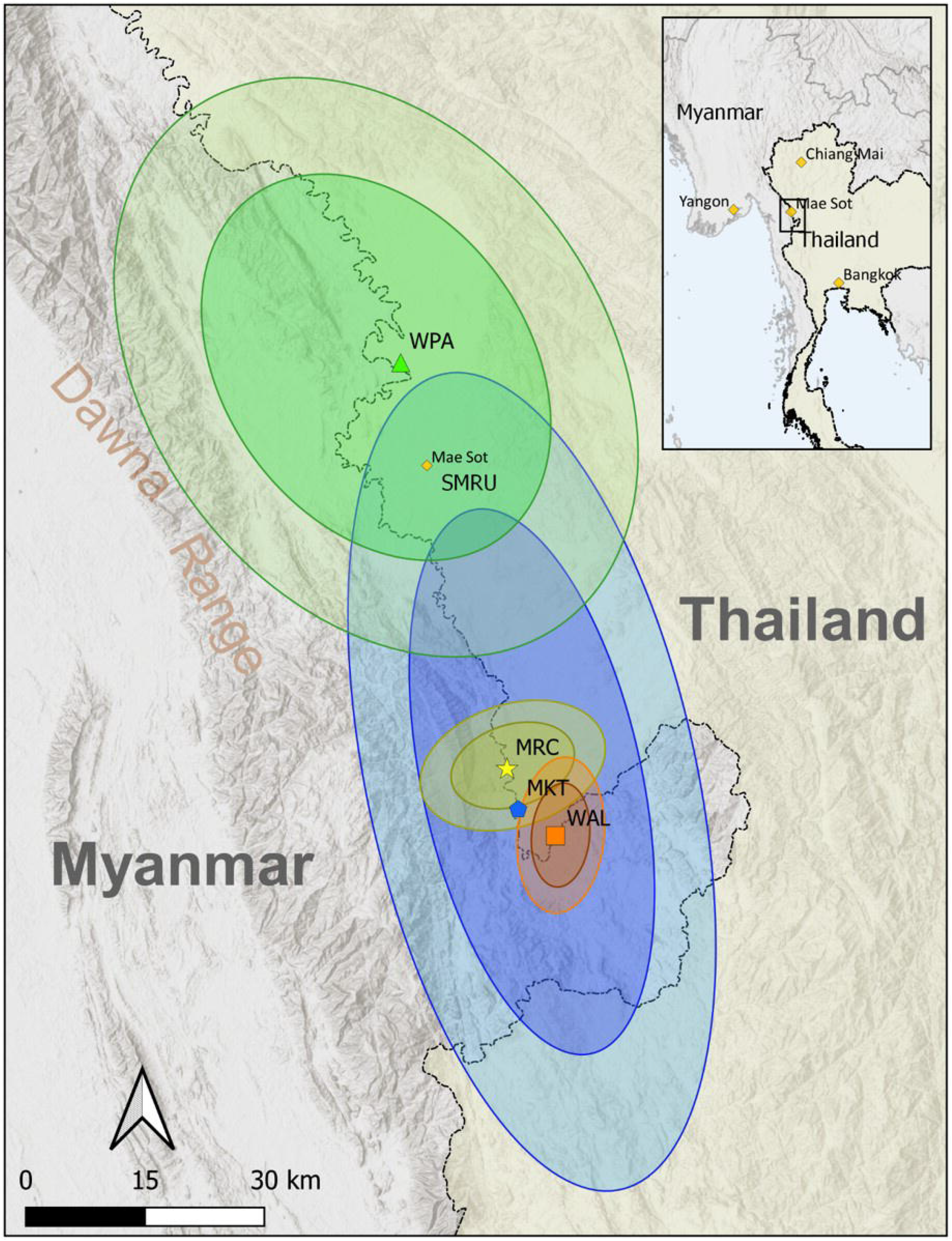
Map of the catchment areas for the four antenatal clinics (ANCs) from 2007-2015. Each of the four clinics is indicated by a different color scheme. The ellipses are standard deviational ellipses, with 2 and 3 standard deviations, explained in detail in the supplementary materials. The darker circle represents roughly 98% of women’s home villages for that specific clinic and the lighter circle representing 99.9% of women’s home villages. WPA (green) and MKT (blue) provided both antenatal care and skilled birth attendants, and MRC (yellow) and WAL (orange) provided antenatal care. WPA and MKT had the largest catchment areas whereas MLC and WAL served a more local population. Maps indicating changes in catchment area over time are presented in Supplementary Figure 2.

Travel distance was associated with maternal health service use across the spectrum of pregnancy care. Travel distance was negatively associated with number of antenatal visits (Supplementary Table 2). For antenatal care use, the 18% of women who began antenatal care in their third trimester also travelled a longer median distance compared to those who began in the first and second trimesters (Table 2). Women with miscarriages travelled farther distances compared to women with normal singleton deliveries (median 5.99km versus 5.24km) (Table 3). Women with pregnancy complications also travelled further compared to those without complications. Women with twins travelled longer median distances compared to women with singleton pregnancies (median 5.99km versus 5.24km) (Table 3), very young mothers (e.g. those aged 13-14) travelled a longer median distance for antenatal care compared to older mothers (Table 4), and women from Myanmar with *P. falciparum* malaria travelled further distances than those with *P. vivax* infections (Table 5, Supplementary Table 3). At the end of the pregnancy spectrum, women who were lost to follow-up and had no record of delivery in a facility with skilled birth services travelled 33% farther compared to women who had normal singleton deliveries in a skilled birth facility (median 6.96km versus 5.24km) (Table 3).

**Table 2:** Distance from home village to health facility by trimester in which patient first presented for antenatal care.

| **Trimester of first attendance** | **min** | **Q1** | **median** | **Q3** | **max** | **count** |
| --- | --- | --- | --- | --- | --- | --- |
| first | 0.00 | 0.89 | 4.72 | 7.52 | 33.82 | 6820 |
| second | 0.00 | 0.89 | 6.31 | 9.35 | 33.82 | 6919 |
| third | 0.00 | 1.97 | 7.10 | 15.44 | 33.82 | 2945 |

**Table 3:** Distance from home village to health facility by pregnancy outcome.

| **Pregnancy Outcome** | **min** | **Q1** | **median** | **Q3** | **max** | **count** |
| --- | --- | --- | --- | --- | --- | --- |
| singleton delivery | 0.00 | 0.89 | 5.24 | 8.05 | 33.82 | 10325 |
| twins | 0.00 | 1.10 | 5.99 | 10.50 | 30.62 | 125 |
| miscarriage | 0.00 | 0.89 | 5.99 | 8.73 | 33.82 | 1179 |
| lost to follow-up | 0.00 | 1.97 | 6.96 | 13.18 | 33.82 | 5079 |

**Table 4:** Distance from home village to health facility by patient age.

| Age Group<br>(years) | min | Q1 | median | Q3 | max | count |
| --- | --- | --- | --- | --- | --- | --- |
| 13thr14 | 0.89 | 2.90 | 7.52 | 16.57 | 24.35 | 11 |
| 15thr19 | 0.00 | 1.10 | 5.99 | 9.35 | 33.82 | 2560 |
| 20thr24 | 0.00 | 0.89 | 5.99 | 9.35 | 33.82 | 4861 |
| 25thr29 | 0.00 | 0.89 | 5.99 | 9.04 | 33.82 | 3835 |
| 30thr34 | 0.00 | 0.89 | 5.99 | 8.87 | 33.82 | 2749 |
| 35thr39 | 0.00 | 0.89 | 5.47 | 8.33 | 33.82 | 1892 |
| 40plus | 0.00 | 0.89 | 5.89 | 9.04 | 33.82 | 800 |

**Table 5:** Distance from health facility by malaria status. Pf = *Plasmodium falciparum*, Pv = *Plasmodium vivax*.

| malaria<br>status | min | Q1 | median | Q3 | max | count |
| --- | --- | --- | --- | --- | --- | --- |
| no malaria | 0.00 | 0.89 | 5.99 | 9.35 | 33.82 | 14305 |
| Pf | 0.00 | 1.10 | 5.99 | 8.87 | 33.82 | 891 |
| Pv | 0.00 | 0.89 | 4.73 | 7.43 | 30.98 | 1881 |
| mixed | 0.00 | 0.89 | 5.78 | 7.59 | 30.62 | 385 |

### Results from the negative binomial regression model

Pregnant women who first presented for care in their third trimester came from 46% farther than those who first presented in their first trimester (DR: 1.46; CI: 1.39 – 1.53; from Table 6), and those with homes in Thailand travelled 63% farther than those with homes in Myanmar to receive care at ANCs (DR: 1.63; CI: 1.57-1.69). Women with *P. falciparum* malaria came from 62% farther away (DR: 1.62; CI: 1.44 – 1.82) than women who never had *P. falciparum* (Table 6), and those with *P. vivax* malaria travelled 23% farther than those without *P. vivax* (DR: 1.23; CI 1.15-1.32).

**Table 6:** Results from a negative binomial regression for predictors of distance to the health facility. The results are given as a ratio of the distances traveled (i.e. the distance ratio (DR)) between a variable and its comparison.

| covariate | count | DR (95% CI) |
| --- | --- | --- |
| *Pregnancy Outcome* |  |  |
| normal singleton delivery | 10325 |  |
| Lost to follow-up | 5079 | 1.45 (1.40 - 1.51) |
| miscarriage | 1179 | 1.23 (1.14 - 1.31) |
| twins delivery | 125 | 1.16 (0.96 - 1.40) |
| *Trimester at first visit to ANC* |  |  |
| 1st trimester presentation | 6787 |  |
| 2nd trimester presentation | 6843 | 1.21 (1.17 - 1.26) |
| 3rd trimester presentation | 2918 | 1.46 (1.39 - 1.53) |
| *Country of home village* |  |  |
| Home village in Myanmar | 8900 |  |
| Home village in Thailand | 6669 | 1.63 (1.57 - 1.69) |
| *Malaria infections during pregnancy* |  |  |
| no *P. falciparum* infection | 15817 |  |
| *P. falciparum* infection | 891 | 1.62 (1.44 - 1.82) |
| No *P. vivax* | 14827 |  |
| *P. vivax* infection | 1881 | 1.23 (1.15 - 1.32) |
| *Health facility where patient received care* |  |  |
| MKT clinic | 4487 |  |
| MLC clinic | 1497 | 0.46 (0.41 - 0.50) |
| WAL clinic | 771 | 0.38 (0.32 - 0.46) |
| WPA clinic | 9953 | 1.03 (0.99 - 1.08) |
| *Patient age at initial ANC visit* |  |  |
| Age 13 through 14 | 11 |  |
| Age 15 through 19 | 2560 | 0.81 (0.45 - 1.45) |
| Age 20 through 24 | 4861 | 0.80 (0.45 - 1.44) |
| Age 25 through 29 | 3835 | 0.78 (0.43 - 1.40) |
| Age 30 through 34 | 2749 | 0.77 (0.43 - 1.39) |
| Age 35 through 39 | 1892 | 0.69 (0.38 - 1.25) |
| Age 40 plus | 800 | 0.71 (0.39 - 1.28) |
| *Patient birth date* |  |  |
| year of birth |  | 1.06 (1.05 - 1.07) |
| *Patient parity* |  |  |
| Parity: 0 | 5803 |  |
| Parity: 1 | 3729 | 0.97 (0.92 - 1.02) |
| Parity: 2-3 | 4552 | 1.00 (0.95 - 1.06) |
| Parity: 4-5 | 1930 | 1.09 (1.01 - 1.17) |
| Parity: 6 - 9 | 659 | 1.15 (1.03 - 1.28) |
| Parity: 10+ | 35 | 1.15 (0.78 - 1.69) |
| *Patient's years lived in their home village at time of initial ANC visit* |  |  |
| Lived in home village for less than 1 year | 4654 |  |
| Lived in home village for 1 through 3 years | 2797 | 1.14 (1.09 - 1.19) |
| Lived in home village for 4 through 9 years | 2524 | 0.97 (0.92 - 1.01) |
| Lived in home village for 10 or more years | 1649 | 1.65 (1.57 - 1.74) |

The regression also indicates that women lost to follow-up travelled 45% farther than women who had normal singleton deliveries (distance ratio (DR): 1.45; CI: 1.40 – 1.51) (Table 6). Women who eventually had a miscarriage travelled 23% farther than those who had normal singleton deliveries (distance ratio: 1.23; CI: 1.14 - 1.31).

Associations between pregnancy outcomes and travel distance were consistent in sensitivity analyses as well (Supplementary Table 3).

## CONCLUSION

These results add to a growing body of literature, mainly gathered in low-income and resource-limited settings that highlight how travel distance limits access to care across the spectrum of pregnancy: from an absence of early antenatal care due to late presentation^23^, as a barrier to treatment in complicated pregnancies such as those with malaria,^7,24^ and as a cause of lost to follow-up in childbirth. ^12,15,25^

This analysis represents the first evidence that travel distance contributes to the high proportion of migrants who are lost to follow-up after enrolling in antenatal care on the Thai/Myanmar border. Given the paucity of any other antenatal care or delivery services for migrants in the rural Thai/Myanmar border region, it is likely that most of those lost to follow-up went without antenatal care and gave birth at home without a skilled attendant. Maternal and neonatal mortality are known to increase when women do not receive antenatal care^3^ or deliver with skilled birth attendants, and although birth outcomes amongst women lost to follow-up was not available, they likely experience a higher risk of morbidity and mortality compared to those who attend ANCs for care.^26^

This is the first evidence for geographic barriers to malaria care in pregnancy in this region, demonstrating that women with *P. falciparum*, and to a lesser extent *P. vivax* malaria, travel longer distances for antenatal care, which may contribute to the high rates of loss to follow-up in women with malaria, noted by Moore et al. 2016.^17^ Stratified analysis suggests that this finding is restricted to those living on the Myanmar side of the border (Supplementary Table 4). Health facilities in Thailand have offered free antimalarials to anyone with a malaria diagnosis for decades, and SMRU has provided subsidized transportation to clinic by care for women living on the Thai side of the border. These differences by country of residence may be accounted for by both the decreased burden of *P. falciparum* malaria in Thailand after decades of concerted public health efforts as well as the increased access to care in Thailand provided by subsidized transportation to ANC. Since 2014 there has been a drastic increase in access to malaria diagnosis and treatment in Karen State as well.^21^

Treatment of malaria in pregnancy has been a major priority for preventing poor pregnancy outcomes worldwide, but the Thai/Myanmar border region faces unique challenges due to high rates of *P. falciparum* multi drug resistance^27^ and the lack of a safe, radical cure that eliminates dormant *P. vivax* from the liver in pregnancy.^28^ Recent initiatives have made progress by bringing curative *P. falciparum* treatments to rural villages^21^, but *P. vivax* now accounts for the majority of malaria infections in this region.^28^ Since the current *P. vivax* treatments indicated in pregnancy only treat the blood stage of the infection, public health campaigns must focus on population-wide *P. vivax* screening and treatment of all non-pregnant residents, which is arguably the most effective tool to decrease the prevalence of *P. vivax* malaria in pregnancy in this region.^2^

This study also identified an association between longer distance travelled and late presentation for antenatal care, indicating limited access to mortality-reducing interventions amongst those living far from health facilities. Increased outreach services to enrol this population in ANC care during the first trimester will broaden access to folic acid supplements to prevent neural tube defects, iron supplements to treat anaemia, and HIV antivirals to prevent maternal-newborn transmission. That 59% of the study population (9761/16548; from Table 2) did not present until after the first trimester of pregnancy also suggests the need for clinical guidelines for antenatal care for those who miss their first trimester antenatal appointments.

Although these findings highlight poor access to care amongst those living far from ANC, they also demonstrate the remarkable resilience of women who overcome significant geographic barriers to attain healthcare. Facing high fevers from malaria and other complications of pregnancy, women often must travel long distances by foot to antenatal clinics and skilled birth services in the Thai-Myanmar border region.

These analyses and data are subject to several limitations. First, there are no officially numbered houses or named streets in the study area, and the addresses used in this analysis correspond to village names. The sensitivity analysis was geared toward testing for the reliability of the geocoding approach and the overarching consistency of the results, regardless of the catchment area used in the analysis, provides confidence in the validity of the results. The study population also includes women whose home location can change seasonally based on employment opportunities, and the addresses used only reflect where women lived at the time of their initial ANC visit. To control for this in the analysis a variable for the duration of time lived at the current address was included.

There are many women who were either too sick to travel, too far away, or without the social and financial resources to arrange for transportation to ANC. Data relied on clinic records and passive detection, leaving pregnant women who never came to a clinic or skilled birth facility unrepresented. This could introduce a selection bias for a healthier patient population, which is supported by studies that document an MMR of 250 (95% confidence interval of 150-430) amongst women attending antenatal care^2^ and 721 in rural villages with limited ANC access (no confidence interval available).^29^

Finally, the model used Euclidian (straight-line) distance as a proxy for geographic access, which may not account for other geographic barriers to care such as the availability of roads, the presence of mountain ranges, rivers, and other geographic barriers, seasonal variations in rains that wash out dirt roads and bridges and make travel arduous or only possible when the water recedes. Travel distance and travel time have been found to closely correlate in other limited-resource settings, but no studies have examined whether that relationship holds in the Thai-Myanmar border region. The straight-line distance represents the easiest possible travel pathway, with reality being that travel is much more difficult.

Though there are limitations to this analysis, there are many strengths as well. It was possible to draw on an incredibly rich, longitudinal cohort of women followed across the spectrum of pregnancy, collected and maintained despite floods and armed conflicts. Although most studies on distance and access to maternal healthcare are survey-based or cross-sectional, this prospectively followed cohort follows women throughout the spectrum of maternity, from initial antenatal presentation up through delivery, and provides rich detail on the effect of geography on healthcare utilization throughout the entirety of pregnancy and across time. Further, until the expansion of malaria services into Kayin State in 2014, such detailed geographic information was almost completely lacking. This analysis builds on the GIS established for the malaria elimination efforts in order to begin to understand the maternal and child health landscape of this region.

The results highlight a consistent problem in this region: poor access to healthcare contributing to loss to follow-up at ANCs in a region with persistently high maternal mortality. Empirical evidence is provided of geographic barriers to antenatal services for pregnant migrant women in Kayin state, Myanmar and Tak province, Thailand. To address these barriers, future ANCs could follow the model of SMRU’s malaria elimination task force, which in 2014 trained a network of rural malaria post workers that substantially reduced the transmission of *P. falciparum* malaria.^21^ Additionally, enhanced outreach services could be used to expand services outside of the walls of existing ANCs and into more rural, hard-to-reach villages. Regardless of the interventions selected, these findings speak to the need for both increased access to antenatal services as well as an enhanced public health surveillance system to proactively monitor health status and provide faster interventions to improve the accessibility of reproductive health care services. These findings can inform efforts to restructure healthcare on both the Thai/Myanmar border and in other settings where patients are lost to follow-up.

At least two further lines of research in this region are recommended. Quality of care may impact travel distances and has recently been shown to have a pronounced effect on decisions of where to seek care in other resource-limited settings.^30^ A more nuanced analysis of distance including quality of care measures would be important. Women in Thailand travelled 63% farther than women in Myanmar, but it remains unknown whether this was related to subsidized transport provided by SMRU, better roads and transportation infrastructure in Thailand, or other factors like availability of other ANC services.

## Data Availability

Access to the data used in this analysis can be requested through the Mahidol-Oxford Tropical Medicine Research Unit data access policy. Both the policy and application form are available at: http://www.tropmedres.ac/data-sharing.

## Acknowledgements

Very many thanks to Rebecca for her help in identifying village locations and matching names. The efforts of the many METF mappers who helped collect the coordinates and place names of the villages throughout Kayin State of Myanmar remain deeply appreciated and hopefully the combined efforts lead us towards a healthier future in the region.

