## Supplementary Files for "Distance matters: barriers to antenatal care and safe childbirth in a migrant population on the Thailand-Myanmar border from 2007-2015, a pregnancy cohort study"

**Supplementary Appendix**

**Appendix Table of Contents**

**Description of analyses**

Estimating patient distance to clinic:	Page 3

Estimating clinic catchment areas (standard deviational ellipses):	Page 3

Sensitivity analysis:	Page 3

**Supplementary Figures**

Figure 1: Histogram of distances to antenatal (ANC) clinics:	Page 4

Figure 2: Maps indicating catchment areas for the clinics over time:	Page 5

**Supplementary Tables**

Table 1: Table of covariates used in formal analysis:	Page 6

Table 2: Summary statistics for number of consultations by distance to clinic:	Page 7

Table 3: Results from sensitivity analysis:	Page 8

Table 4: Formal analysis results stratified by nation of origin (Thailand or Myanmar):	Page 9

Table 5: Summary statistics for distance to clinic by year and clinic:	Page 11

1. **Estimating distance travelled to clinic.**

We created a pairwise Euclidian (i.e. straight line) distance matrix between each patient’s village and the ANC clinic they attended during pregnancy. Using this distance matrix, we created summary statistics and performed univariate analyses that assessed differences in distance to clinic by variables listed in the patient records. Variables included year of childbirth, parity, age, malaria infection status, and pregnancy outcome (singleton delivery, twins, lost to follow-up, or miscarriage).

2. **Estimating clinic catchment areas.**

We created visual representations of the spatial distribution of distance from village to clinic using standard deviational ellipses (SDEs). SDEs measure 2-dimensional spread (the standard deviation) along an X- and Y-axis from the mean center of a set of points. The Y-axis is then rotated until the sum of the squares of the distances between points (village locations) and axes are minimized. The angle is defined as:

$$\theta=arctan\left\{ \frac{\left[ \sum_{i=1}^{n} \left( x_{i}-X_{MC} \right)^{2}-\sum_{i=1}^{n} \left( y_{i}-Y_{MC} \right)^{2} \right]+\sqrt{\left[ \left\{ \left( \sum_{i=1}^{n} \left( x_{i}-X_{MC} \right)^{2}-\sum_{i=1}^{n} \left( y_{i}-Y_{MC} \right)^{2} \right) \right\}^{2}+4\left\{ \sum_{i=1}^{n} \left( x_{i}-X_{MC} \right)\left( y_{i}-Y_{MC} \right)^{2} \right\} \right]}}{2\sum_{i=1}^{n} \left( x_{i}-X_{MC} \right)\left( y_{i}-Y_{MC} \right)} \right\}$$

The standard deviation is then calculated along both the shifted X- and Y-axes:

$$s_{X}= \sqrt{\frac{\sum_{i=1}^{n} \left[ \left( x_{i}-X_{MC} \right)\cos\theta-\left( y_{i}-Y_{MC} \right)\sin\theta\right]}{n}}$$

$$s_{Y}=\sqrt{\frac{\sum_{i=1}^{n} \left[ \left( x_{i}-X_{MC} \right)\sin\theta+\left( y_{i}-Y_{MC} \right)\cos\theta\right]}{n}}$$

.

The output of these metrics is frequently visualized as an ellipse layer on a map; with 1, 2, or 3 standard deviations that roughly correspond to 63, 98, or 99 % of all geographic points, respectively, falling within the ellipse. Spatial point patterns that are isotropic will result in an SDE that is circular whereas anisotropic distributions will be more elliptic.

3. **Sensitivity analysis**

Since some error is expected in the geocoding process, a sensitivity analysis was conducted on the regression based on estimated distances to the nearest maternal health facility. This was based on the hypothesis that geocoding was most accurate with villages nearest to ANCs, with which the health facility staff maintaining medical records are most familiar, and less accurate farther from ANCs. The regression was re-run amongst subgroups of women with home villages within 30km, 20km, 10km, and 5km of ANCs. Model results were compared across all distance restrictions (Supplementary Table 2).

**SUPPLEMENTARY FIGURES**

**Supplementary Figure 1**: Histogram showing distribution of distances to ANCs (antenatal clinics) for all patients. The unit of measure for the x-axis is km, and the y-axis is number of individual patients between 2007-2015 at all four ANC clinics for a given distance from their closest clinic.

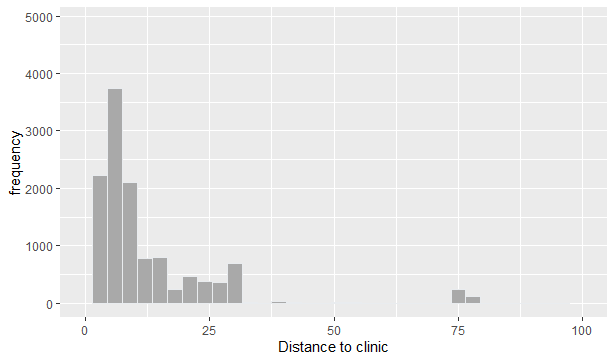

**Supplementary Figure 2:** Maps indicating catchment areas for the clinics over time. Each of the four clinics is indicated by a different color scheme. The ellipses are standard deviational ellipses (with 2 and 3 standard deviations), with the darker circle representing roughly 98% of women’s home villages for that specific clinic and the lighter circle representing 99.9% of women’s home villages. WPA (green) and MKT (blue) provided both antenatal care and skilled birth attendants, and MRC (yellow) and WAL (orange) provided antenatal care.

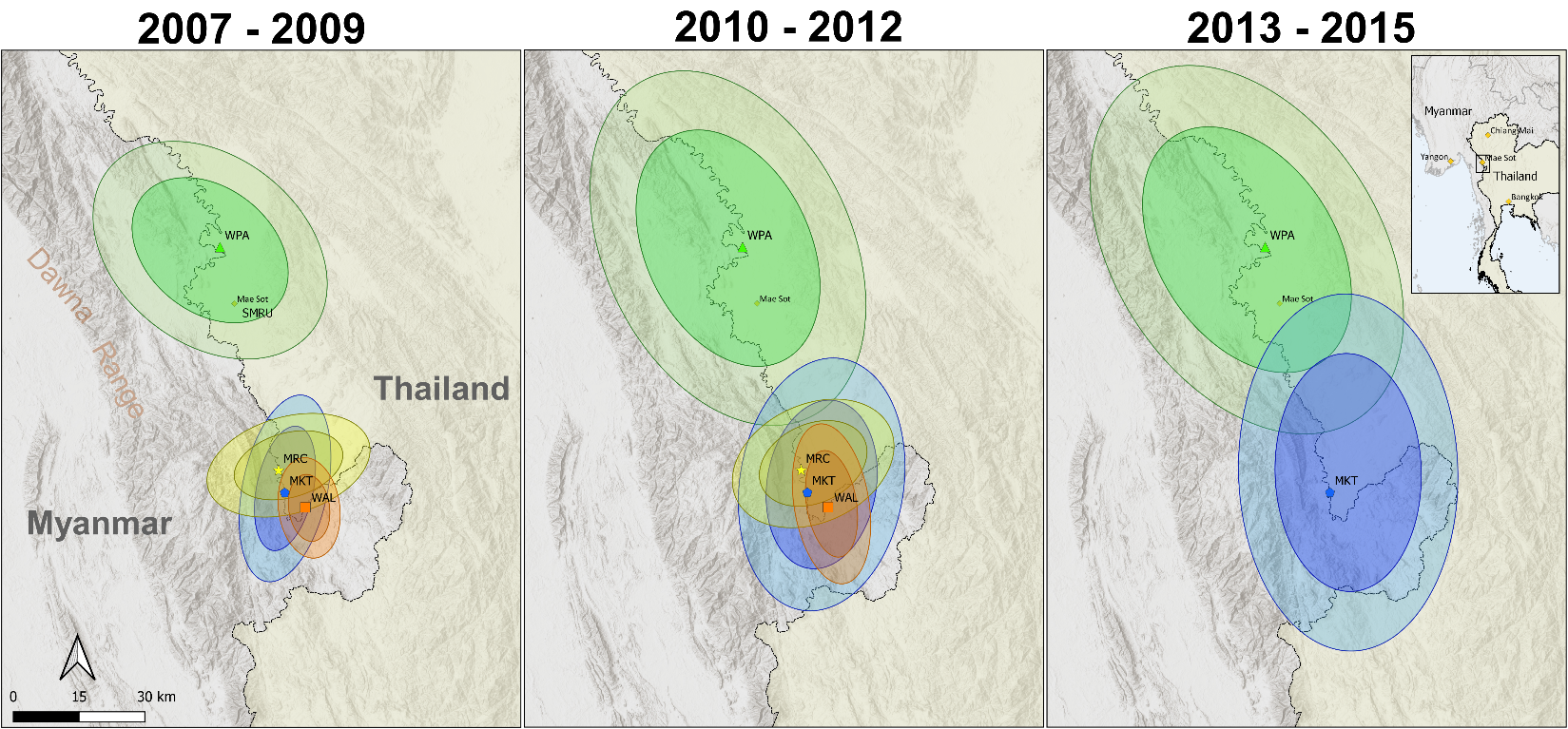

**SUPPLEMENTARY TABLES**

**Supplementary Table 1:** List and explanation of covariates in the negative binomial regression models

| **Covariate** | **specification** | **categories** |
| --- | --- | --- |
| *pregnancy outcome* | categorical | normal singleton, lost, miscarriage, or twins |
| *clinic* | categorical | MKT, MLC, WAL, or WPA |
| *P. falciparum malaria* | categorical/binary | yes = 1, no = 0 |
| *P. vivax malaria* | categorical/binary | yes = 1, no = 0 |
| *trimester at which pregnant women first presented at the ANC* | ordinal | 1st, 2nd, or 3rd |
| *which nation the person lives in* | categorical | Myanmar or Thailand |
| *age* | ordinal | 13 through 14, 15 through 19, 20 through 24, 25 through 29, 30 through 34, 35 through 39, 40 plus |
| *year of pregnancy outcome* | continuous | from 2007 through 2015 |
| *parity* | ordinal | parity 0, parity 1, parity of 2-3, parity of 4-5, parity of 6 - 9, parity of 10+ |
| *years living at current residence* | ordinal | less than 1 year, 1 through 3 years, 4 through 9 years, 10 or more years |

**Supplementary Table 2**: Summary statistics for number of consultations (NOC) by distance from home village to clinic. NOC was also strongly correlated with parity and pregnancy outcomes (i.e. those with miscarriages had fewer NOC than those with normal deliveries).

| **distance** | **min** | **Q1** | **median** | **mean** | **Q3** | **max** | **count** |
| --- | --- | --- | --- | --- | --- | --- | --- |
| same town | 1 | 4 | 10 | 11.17 | 17 | 37 | 1638 |
| <5km | 1 | 4 | 8 | 9.65 | 14 | 37 | 6276 |
| 6 - 10km | 1 | 3 | 7 | 8.08 | 12 | 35 | 5060 |
| 11 - 15km | 1 | 2 | 4 | 5.35 | 8 | 35 | 1386 |
| 16 - 20km | 1 | 1 | 3 | 4.71 | 7 | 19 | 601 |
| 21 - 25km | 1 | 1 | 3 | 4.19 | 6 | 32 | 675 |
| 26 - 30km | 1 | 1 | 3 | 4.69 | 6 | 33 | 652 |
| 31 - 35km | 1 | 2 | 5 | 6.81 | 10 | 34 | 421 |
| >35km | 1 | 1.5 | 3 | 3.52 | 4 | 11 | 34 |

**Supplementary Table 3**: Results from a sensitivity analysis on the distance to clinic. The model was re-run using different cutoffs for distance to clinic, based on our hypothesis that geotagging was more accurate for villages nearer to the clinics. Most results remain consistent until we limit the analysis to a small radius around the clinic (5km or less).

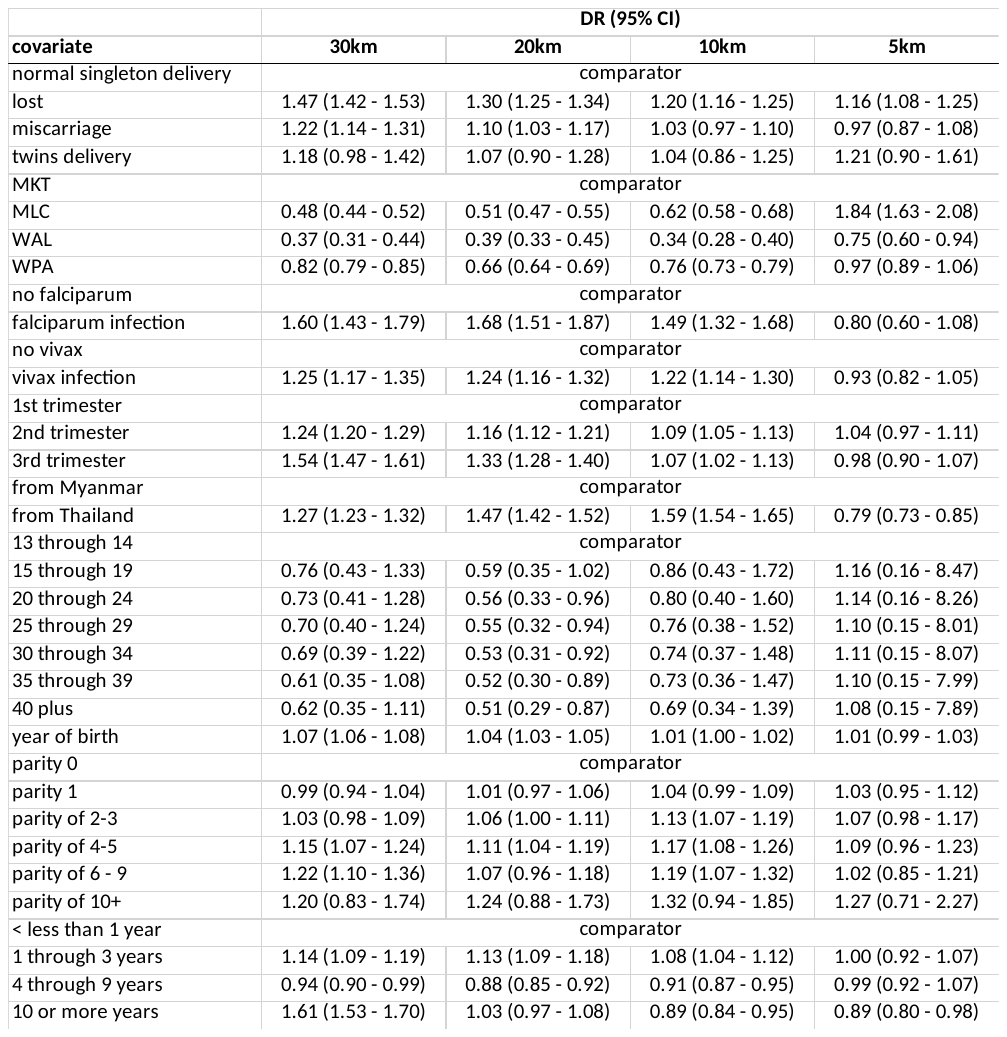

The sensitivity analysis also revealed that women who eventually had miscarriages travelled a greater distance in the 30, 20, and 10km subgroups, but in the 5km subgroup women with and without miscarriages had no travel distance differences. The sensitivity analysis suggested that women with pregnancies complicated by *P. falciparum* and *P. vivax* infections travelled a greater distance in the 30, 20, and 10km subgroups. In the 5km subgroup, however, women with and without malaria infections had no travel distance differences.

Women lost to follow-up travelled significantly farther in the 30, 20, 10, and 5km subgroups (Supplementary table 1). That the magnitude and direction of this effect was consistent across all four subgroups increases confidence that geocoding accurately labelled women’s villages out to 30km away from the health facility. If the hypothesis that geocoding accuracy is inversely proportional to travel distance had been true, the expected findings would have dissipated in subgroups that included women living farther from the health facility.

**Supplementary Table 4:** Results from a negative binomial regression for predictors of distance to the clinic, stratified by nation of origin (Myanmar and Thailand).

| **covariate** | **Myanmar** | | **Thailand** | |
| --- | --- | --- | --- | --- |
|  | **count** | **DR (95% CI)** | **count** | **DR (95% CI)** |
| normal singleton delivery | 5701 |  | 3879 |  |
| lost | 2423 | **1.69 (1.60 - 1.78)** | 2308 | **1.18 (1.12 - 1.24)** |
| miscarriage | 705 | **1.30 (1.19 - 1.43)** | 432 | 1.09 (0.99 - 1.20) |
| twins delivery | 71 | **1.30 (1.01 - 1.68)** | 50 | 1.06 (0.82 - 1.38) |
| MKT | 1727 |  | 2760 |  |
| MLC | 274 | **0.45 (0.37 - 0.56)** | 927 | **0.47 (0.42 - 0.52)** |
| WAL | 431 | **0.28 (0.22 - 0.35)** | 158 | **0.67 (0.51 - 0.89)** |
| WPA | 6468 | **1.07 (1.01 - 1.14)** | 2824 | **0.93 (0.89 - 0.98)** |
| no falciparum | 8287 |  | 6584 |  |
| falciparum infection | 613 | **1.67 (1.47 - 1.91)** | 85 | 1.01 (0.73 - 1.41) |
| no vivax | 7660 |  | 6333 |  |
| vivax infection | 1240 | **1.35 (1.24 - 1.47)** | 336 | 0.94 (0.82 - 1.08) |
| 1st trimester | 3601 |  | 2552 |  |
| 2nd trimester | 3607 | **1.17 (1.11 - 1.24)** | 2829 | **1.22 (1.16 - 1.29)** |
| 3rd trimester | 1616 | **1.40 (1.31 - 1.50)** | 1238 | **1.40 (1.31 - 1.49)** |
| 13 through 14 | 7 |  | 3 |  |
| 15 through 19 | 1383 | 0.84 (0.39 - 1.78) | 1004 | 0.62 (0.26 - 1.51) |
| 20 through 24 | 2530 | 0.76 (0.36 - 1.61) | 2022 | 0.69 (0.29 - 1.67) |
| 25 through 29 | 2028 | 0.70 (0.33 - 1.50) | 1536 | 0.70 (0.29 - 1.70) |
| 30 through 34 | 1468 | 0.68 (0.32 - 1.46) | 1073 | 0.71 (0.29 - 1.73) |
| 35 through 39 | 1057 | 0.56 (0.26 - 1.20) | 704 | 0.73 (0.30 - 1.76) |
| 40 plus | 427 | 0.55 (0.25 - 1.17) | 327 | 0.79 (0.32 - 1.92) |
| year of birth |  | **1.06 (1.05 - 1.08)** |  | **1.05 (1.03 - 1.06)** |
| parity 0 | 2926 |  | 2556 |  |
| parity 1 | 2017 | 1.04 (0.97 - 1.11) | 1469 | 0.92 (0.86 - 0.98) |
| parity of 2-3 | 2462 | **1.12 (1.04 - 1.21)** | 1723 | **0.90 (0.84 - 0.97)** |
| parity of 4-5 | 1072 | **1.30 (1.17 - 1.45)** | 694 | **0.90 (0.81 - 0.99)** |
| parity of 6 - 9 | 401 | **1.50 (1.30 - 1.72)** | 217 | **0.80 (0.69 - 0.93)** |
| parity of 10+ | 22 | **1.67 (1.01 - 2.75)** | 10 | 0.61 (0.34 - 1.10) |
| < less than 1 year | 2210 |  | 2444 |  |
| 1 through 3 years | 1484 | **1.22 (1.14 - 1.30)** | 1313 | 1.05 (0.99 - 1.11) |
| 4 through 9 years | 1569 | 0.95 (0.89 - 1.01) | 954 | 1.00 (0.94 - 1.06) |
| 10 or more years | 1260 | **1.99 (1.86 - 2.13)** | 389 | 0.79 (0.72 - 0.87) |

The model for patients coming from Myanmar largely corresponds to the full model. Those who were lost to follow up were coming from farther away (69% farther) than patients who had normal singleton deliveries. Those who had a P. falciparum infection during their pregnancy were coming from 67% farther away and those with P. vivax infections from 35% farther away. Those who presented for the first time at the ANC in their 3^rd^ trimester were coming from 40% farther away than those who presented in the 1^st^ trimester.

The model for patients coming from Thailand differed in that there was no statistically significant difference in distances for patients based on malaria infections. Still, those who were lost to follow up were coming from farther (18% farther) away than those with normal singleton deliveries (though the difference in distance was smaller than for women coming from Myanmar). As with the other models, those who first presented later in their pregnancy were coming from farther away than women who presented in their first trimester.

**Supplementary Table 5:** Summary statistics for distance travelled to clinic (median (mean) in kilometers) by year and number of patients. Both MLC and WAL closed during the study period and many patients from these clinics began attending nearby MKT (resulting in an increase in no. of patients and an increase in the median (mean) distance travelled to MKT clinic).

|  | **MLC** | | **MKT** | | | **WAL** | | **WPA** | |
| --- | --- | --- | --- | --- | --- | --- | --- | --- | --- |
| **Year** | **median (mean) distance** | **no. patients** | | **median (mean) distance** | **no. patients** | **median (mean) distance** | **no. patients** | **median (mean) distance** | **no. patients** |
| 2007 | 4.32 (4.20) | 330 | 5.78 (6.71) | | 17 | 0.00 (1.36) | 189 | 0.89 (3.98) | 743 |
| 2008 | 4.32 (3.94) | 258 | 2.17 (3.49) | | 162 | 0.00 (1.75) | 224 | 1.97 (4.66) | 881 |
| 2009 | 4.32 (3.74) | 258 | 2.17 (4.05) | | 210 | 0.00 (1.62) | 180 | 4.00 (6.33) | 1196 |
| 2010 | 4.32 (4.58) | 297 | 5.78 (5.51) | | 263 | 0.00 (2.77) | 173 | 6.58 (8.05) | 1217 |
| 2011 | 4.32 (4.72) | 269 | 5.99 (7.04) | | 434 | 0.00 (0.53) | 5 | 6.58 (8.40) | 1225 |
| 2012 | 4.32 (3.66) | 85 | 6.31 (8.25) | | 588 | clinic closed | | 6.42 (8.05) | 1151 |
| 2013 | clinic closed | | 7.52 (9.94) | | 908 |  |  | 6.88 (8.75) | 1124 |
| 2014 |  |  | 10.02 (11.00) | | 956 |  |  | 6.88 (9.19) | 1198 |
| 2015 |  |  | 10.50 (11.53) | | 949 |  |  | 6.88 (9.51) | 1218 |
